# CHANGES IN INCIDENCE AND SEROTYPE DISTRIBUTION OF PEDIATRIC INVASIVE PNEUMOCOCCAL DISEASE AFTER THE INTRODUCTION OF 15-VALENT PNEUMOCOCCAL CONJUGATE VACCINE IN CATALONIA, SPAIN. A MULTICENTER SURVEILLANCE STUDY

**DOI:** 10.64898/2026.02.11.26346066

**Authors:** C Muñoz-Almagro, M Cisneros, C Alcaraz, S Broner, F Moraga-Llop, A Rosell, A Diaz-Conradi, P Brotons, D Henares, G Gonzalez-Comino, B Viñado, F Gómez-Bertomeu, C Marco, S Gonzalez-Peris, J Llaberia, C Izquierdo, J Galvez, A Perez-Argüello, R Varo, J Iglesies, C Esteva, M Armas, M Blanco-Fuertes, N Torrellas, MO Perez, IT Valle, M Navarro, A Rivera, M Colomer, L Solaz, M Mico, JJ Garcia-Garcia, A Dominguez, MF de Sevilla, P Ciruela, Catalan Study Group of Invasive Pneumococcal Disease

## Abstract

**Background:** Serotype 3 (S3) has remained a major cause of invasive pneumococcal disease (IPD) despite its inclusion in 13-valent pneumococcal conjugate vaccine (PCV). In October 2023, a 15-valent PCV (PCV15) including S3 was introduced into the Catalan universal childhood immunization program.

**Methods:** We conducted a retrospective pre-post surveillance study to compare pediatric IPD incidence in Catalonia during a pre-PCV15 period (October 1, 2022-September 30, 2023) and two post-PCV15 periods (October 1, 2023-September 30, 2024, and October 1, 2024-September 30, 2025). All IPD episodes in children <18 years attended in 34 hospitals were included. IPD was defined as detection of *S. pneumoniae* in a sterile site by culture or PCR.

**Results:** 323 IPD episodes were identified in 319 children (mean age, 4.5 years). Overall IPD incidence declined from 13.0 to 9.4 episodes per 100,000 children in the first post-PCV15 period compared with the pre-PCV15 period (28% reduction; *p*=0.02), but returned to baseline in the second post-PCV15 period. S3-IPD incidence decreased significantly from 4.1 to 1.6 episodes per 100,000 (60% reduction; *p*=0.001) in the first post-PCV15 period and remained lower in the second period: 2.3 episodes per 100,000 (42% reduction compared with baseline; *p*=0.04). In contrast, IPD incidence caused by PCV7 serotypes increased from 0.3 in the pre-PCV15 and first post-PCV15 period to 2.7 episodes per 100,000 in the second post-PCV15 period (690% increase; *p*<0.001).

**Conclusion:** PCV15 introduction was associated with a sustained reduction in S3-IPD over two years. However, a marked increase in PCV7 serotypes offset overall gains in IPD incidence.

**SUMMARY:** PCV15 introduction in Catalonia achieved sustained reduction in serotype 3 invasive pneumococcal disease over two years, but a marked increase in PCV7 serotypes offset the overall disease reduction in the second post-vaccination year.

## INTRODUCTION

Invasive pneumococcal disease (IPD) is a major public health problem caused by *Streptococcus pneumoniae*.[1]. This pathobiont bacterium commonly colonizes the human nasopharynx asymptomatically, but it can cause diseases ranging from mild infection to life-threatening IPD [2]. There are more than 107 serotypes of *S. pneumoniae*, and each has a different ability to cause disease [3]. Pneumococcal conjugate vaccines (PCVs) are highly effective against IPD caused by vaccine-covered serotypes [4]. Additionally, PCVs prevent nasopharyngeal colonization by vaccine serotypes, which is the first step in the development of IPD. Furthermore, these vaccines have been shown to limit pneumococcal transmission, offering both direct and indirect (herd) protection [5].Through vaccination, the circulation of pneumococcal vaccine-serotypes is decreased, and vaccinated infants indirectly protect individuals who are not part of the vaccine’s target population.

In the early 2000s, a 7-valent pneumococcal conjugate vaccine (PCV7), covering serotypes 4, 6B, 9V, 14, 18C, 19F, and 23F, was licensed for use in infants and young children, first in the United States and later in Europe. A decline of disease caused by vaccine serotypes was reported early [6]. Overall, this vaccine proved to be highly effective against vaccine serotypes worldwide [7], although the emergence of non-vaccine serotypes limited its impact [8].

Two additional pneumococcal conjugate vaccines targeting emerging serotypes were licensed between 2009 and 2010 in Europe: the 10-valent pneumococcal conjugate vaccine (PCV10), which included serotypes 1, 5, and 7F, in addition to the PCV7 serotypes, and the 13-valent vaccine (PCV13), which targets serotypes 3, 6A, and 19A, in addition to the PCV10 serotypes. After its licensure, the Vaccination Advisory Committee of the Spanish Association of Pediatrics recommended the routine administration of PCV13. In July 2016, PCV13 was incorporated into the childhood immunization program in Catalonia, a region located in northeastern Spain, and was provided free of charge to young children. Early studies in this region reported good effectiveness of at least one dose of PCV13 in preventing IPD caused by vaccine serotypes in children aged 7–59 months, except for serotype 3 [9]. Reduced PCV13 effectiveness against serotype 3 was also documented in the United Kingdom [10].

In subsequent years, several studies consistently reported small effects of PCV13 on the burden of IPD caused by serotype 3 [11]. A multicenter European study found that serotype 3 accounted for 9% of all IPD cases and was the most common serotype causing IPD in children <5 years of age in 2018 [12]. Similarly, in our region, serotype 3 ranked third among IPD isolates in children < 5 years and the first among adults > 65 years, even after the implementation of PCV13 [13]. Persistence of this serotype has been linked to vaccine failures [14] and severe clinical outcomes, including necrotizing pneumonia [15].

In response to ongoing epidemiological challenges two new pneumococcal conjugate vaccines were approved by the European Commission in 2022: the 15□valent pneumococcal conjugate vaccine (PCV15) for children, which covers PCV13 serotypes and adds 22F and 33F serotypes, [16] and the 20□valent pneumococcal conjugate vaccine (PCV20) for adults older than 18 years, which targets PCV15 serotypes and adds 8, 10A, 11A, 12F, and 15B serotypes [17]. The European Commission approved PCV20 for children in 2024 [18].

In September 2022, the Catalan Department of Health included PCV20 in the universal immunization program for adults aged 65 years or older [19] and in October 2023, PCV13 was replaced by PCV15 in the universal immunization program for children, following a 2□+□1 schedule at 2, 4, and 11 months of age [20]. While uptake of PCV20 has been estimated to be low in the adult target population, high vaccine coverage with PCV15 has been achieved in infants. According to the Vaccination Information System of the Spanish Ministry of Health (SIVAMIN), PCV15 coverage in Catalonia in 2024 was 96.17 % for at least two doses [21]. Of note, in 2025, most Spanish autonomous communities incorporated PCV20 into their routine childhood immunization programs, except for Catalonia, Navarra, Madrid, and the Basque Country, which adopted PCV15 [22].

In October 2023, coinciding with PCV15 introduction for young children, universal immunization with the monoclonal antibody nirsevimab for all children under 6 months of age was started in the majority of Spanish regions to prevent bronchiolitis caused by respiratory syncytial virus (RSV). It has been estimated that an average of 92% of newborns were immunized with nirsevimab in Spain during 2023-2024 [23].

Given the recent implementation of PCV15 in the pediatric population, the current dynamics of IPD in Catalonia remain largely undefined. This study aims to assess the impact of PCV15 introduction on IPD cumulative incidence and serotype distribution among children in Catalonia.

## PATIENTS AND METHODS

### Study design

A retrospective population-based pre-post surveillance study was conducted to compare the cumulative incidence of pediatric IPD in Catalonia in a pre-PCV15 basal period (October 1, 2022 - September 30, 2023) *versus* two post-PCV15 periods including period 1 (October 1, 2023 -September 30, 2024) and period 2 (October 1, 2024 - September 30, 2025). The study included all pediatric IPD episodes detected in the study setting.

### Study Setting

All pediatric IPD patients attended in hospitals across Catalonia were included. During the study period, 34 hospitals systematically submitted pneumococcal isolates or pneumococcal DNA extracts detected in invasive clinical samples to the regional reference laboratory responsible for molecular surveillance of IPD. This reference laboratory is located at Sant Joan de Déu University Hospital (HSJD) in Barcelona. Although submission of samples for molecular characterization is not mandatory for hospitals in Catalonia, these participating centers consistently referred all identified invasive samples to the reference surveillance laboratory throughout the study period (the list of participating hospitals is provided in Table S1).

During the study period, the 34 participant hospitals provided care to approximately 70% of children younger than under 2□years of age residing in Catalonia, 67% of children in the age range 2-4 and 65% of children in the age range 5-18 years (data of reference pediatric population residing in the catchment area of the participant hospitals is provided in Supplementary Table S2)

#### Definition of invasive pneumococcal disease

IPD was defined as the presence of clinical findings of infection confirmed by isolation or DNA detection off *S. pneumoniae* by real-time PCR in a normally sterile sample. IPD was classified according to the International Classification of Diseases, 11th Revision [24] code specific for diseases caused by *S. pneumoniae,* including meningitis, pneumonia, parapneumonic empyema, occult bacteremia, sepsis, arthritis, peritonitis, and endophthalmitis. Recurrent IPD was defined as two or more episodes in the same individual occurring at least one□month apart.

### Data sources

The pediatric study population under surveillance was inferred from the total Catalan pediatric population, which is recorded annually and available from the “Padró municipal d’habitants” (PMH) public database website (www.idescat.cat). Similarly, the number of pediatric hospital admissions to the hospitals participating in this multicenter study was inferred from the total pediatric hospitalizations across Catalonia, which is registered and publicly available with yearly updates on a per-age and per-hospital basis on the Minimum Basic Data Set (CMBD) public database website (accessible at: https://catsalut.gencat.cat).

### Study variables

The following variables were considered for the study: patient age group (< 2 years, 2-4 years, 5-17 years), sex, IPD clinical manifestation (bacteremia/sepsis, meningitis, pneumonia, complicated pneumonia, and others), detection technique for *S. pneumoniae* (culture/PCR or PCR alone), and serotype vaccine coverage (PCV7-covered, PCV13-covered, PCV15-covered, PCV20-covered and non-PCV20-covered). Outcomes included pre- and post-PCV15 cumulative incidence by age group and serotype vaccine coverage, considering the pre-PCV15 period as baseline.

IPD cumulative incidence was calculated as the relation between the number of pediatric hospitalizations in the participant hospitals in the period and age group under consideration (numerator) and the reference pediatric population resident in the catchment area of the participant hospitals (denominator) for that period and age group. To estimate the per-period and per-age group pediatric population residing in the catchment area of the participant hospitals, a coefficient rating the number of pediatric hospitalizations in the participant hospitals in relation to the total number of pediatric hospitalizations in Catalonia (both data retrieved from the CMBD database) was applied to the corresponding Catalan population (retrieved from the PMH database).

### Laboratory methods

Microbiological identification of IPD was conducted at the hospitals of origin through the implementation of standard microbiological techniques (including Gram staining, culturing requirements, colony morphology, optochine sensitivity testing, and bile solubility testing for isolates) and detection of *lytA* gene if available in their centers. Following presumptive confirmation, the strains or positive *lytA* DNA extracts were submitted to the Molecular Microbiology Department of HSJD for confirmation of IPD and capsular typing. *S. pneumoniae* DNA was confirmed through amplification of the autolysin-encoding gene (*lytA*) and the Wzg-encoding gene (*cpsA*) according to criteria published elsewhere [25]. For DNA extracts, only those samples that were positive for the *lytA* and *cpsA* genes by real-time PCR were included.

### Microbiological identification and, capsular typing

Strains were confirmed and typed by Whole Genome Sequencing by using Illumina sequencing and the *Pathogen watch* software, as previously described [26].

Culture-negative, but PCR-positive samples were confirmed and typed using two methods, based on *S. pneumoniae* DNA quantity. For low DNA amounts (detection of *LytA* gene DNA with the cycle threshold [CT] of >30 cycles),, real-time multiplex PCR technique targeting the *cpsA* gene for detection of all pneumococcal capsular types and differentiating serotypes 1, 3, 4, 5, 6A/C, 6B/D, 7F/A, 8, 9V/A/N/L, 14, 15B/C, 18C/B, 19A, 19F/B/C, 23A, and 23F was used . For high DNA amounts (PCR-positive samples with CT of ≤30 cycles), sequential multiplex PCR combined with fragment analysis and automated fluorescent capillary electrophoresis targeting the *cpsA* gene for detection of all capsular types and to differentiate serotypes [1, 2, 3, 4, 5, 6A/6B, 6C, 7F/7A, 7C/(7B/40), 8, 9V/9A, 9N/9L, 10A, 10F/(10C/33C), 11A/11D/11F, 12F/(12A/44/46), 13, 14, 15A/15F, 15B/15C, 16F, 17F, 18/(18A/18B/18C/18F), 19A, 19F, 20, 21, 22F/22A, 23A, 23B, 23F, 24/(24A/24B/24F), 31, 33F/(33A/37), 34, 35A/(35C/42), 35B, 35F/47F, 38/25F, and 39.] was performed [25].

Serotypes were classified into PCV7 serotypes (serotypes included in the 7-valent vaccine), PCV13 serotypes (those added to the 13-valent vaccine), PCV15 serotypes (those added to the 15-valent vaccine), PCV20 serotypes (those added to the 20-valent vaccine), and serotype 3. Serotype 3 was analyzed separately due to its reduced vaccine effectiveness [14]. Of, note, serotype 15B is included in PCV20, and serotype 15C is not, although immunogenicity studies suggest cross-protection from serotype 15B. Since our molecular methods cannot differentiate between serotypes 15B and 15C, we report them collectively as 15B/C and classify them as PCV20 serotypes. Similarly, serotype 33F is included in PCV15, while serotype 37 is not. Given that our molecular methods do not distinguish between serotypes 33F and 37, we report them as 33F and consider them included in the PCV15. Additionally, serotype 12F is included in PCV20, whereas serotypes 12A, 44, and 46 are not. Due to limitations of our molecular methods in distinguishing among these serotypes, we report them as 12F and consider them as PCV20 serotypes.

### Statistical analysis

Proportions of categorical variables were compared using the chi-square or Fisher’s exact test, with Fisher’s test applied when ≥25% of cells had expected frequencies <5. Global cumulative incidence of IPD during the pre- and both post-PCV15 periods was calculated by dividing the number of IPD cases by the total catchment area population, assuming a constant pediatric population throughout each period. The cumulative incidence and changes in serotype distribution were compared across periods using the chi-square or Fisher’s test, and were reported with 95% confidence interval (CI). Two-sided *p*-values of <0.05 were considered to be statistically significant. Statistical analyses were conducted using SPSS for Windows, version 29.0 (SPSS) and R software version 4.4.2 .

### Ethics statement

Molecular surveillance of IPD was approved by the Ethics Committee of HSJD when this hospital was designated as the support laboratory of the Public Health Agency of Catalonia in 2009. No informed consent was requested for this study, as it was based on molecular epidemiology surveillance, and the samples were duly anonymized.

## RESULTS

A total of 323 IPD episodes in 319 children were detected, including 178 (55,8%) male and 141 (44,2%) female, with a mean age of 4.5 years (standard deviation SD=4.20). One-hundred and twenty-one episodes occurred in the pre-PCV15 period, 85 in the first post-PVC15 period, and 117 in the second post-PCV15 period. Two hundred and twenty-seven (70.3%) of 323 episodes were detected by culture/Real-Time PCR and 96 (29.7%) only by real-time PCR.

The main clinical manifestation of IPD was pneumonia in 187 (57.9%) of episodes. Among pneumonia episodes, 29.4% were complicated pneumonia (n= 55). (Table 1).

**Table 1:** Epidemiological and clinical characteristics of study population.

|  | Total |  | Before PCV15 |  | 1 year after |  | 2 years after |  | p-value |
| --- | --- | --- | --- | --- | --- | --- | --- | --- | --- |
| Variables | (n=323) |  | (n=121) |  | PCV15 (n=85) |  | PCV15 (n=117) |  |  |
|  | N | (%) | N | (%) | N | (%) | N | (%) |  |
| Age (mean + IQR) |  |  |  |  |  |  |  |  |  |
| < 2 years | 112 | (34.6%) | 38 | (31.4%) | 30 | (35.3%) | 44 | (37.6%) | 0.07 |
| 2-4 years | 115 | (35.5%) | 54 | (44.6%) | 23 | (27.1%) | 38 | (32.5%) |  |
| 5-17 years | 96 | (29.6%) | 29 | (24.0%) | 32 | (37.6%) | 35 | (29.9%) |  |
| Gender |  |  |  |  |  |  |  |  |  |
| Male | 180 | (55.7%) | 83 | (68.6%) | 45 | (52.9%) | 52 | (44.4%) | <0.001 |
| Female | 143 | (44.3%) | 38 | (31.4%) | 40 | (47.1%) | 65 | (55.6%) |  |
| Clinical manifesta-<br>tion |  |  |  |  |  |  |  |  |  |
| Bacteremia | 101 | (31.3%) | 36 | (29.8%) | 25 | (29.4%) | 40 | (34.2%) | 0.508 |
| Pneumonia | 187 | (57.9%) | 74 | (60.2%) | 46 | (54.1%) | 67 | (57.3%) |  |
| *Non-complicated* | 132 | (70.6%) | 48 | (64.9%) | 34 | (73.9%) | 50 | (74.6%) |  |
| *Complicated* | 55 | (29.4%) | 26 | (35.1%) | 12 | (26.1%) | 17 | (25.4%) |  |
| Meningitis | 28 | (8.7%) | 9 | (7.4%) | 12 | (14.1%) | 7 | (6.0%) |  |
| Others | 7 | (2.2%) | 2 | (1.6%) | 2 | (2.4%) | 3 | (2.6%) |  |
| Microbiological<br>test/ techniques |  |  |  |  |  |  |  |  |  |
| Culture | 227 | (70.3%) | 82 | (67.8%) | 68 | (80.0%) | 77 | (65.8%) | 0.07 |
| Only PCR | 96 | (29.7%) | 39 | (32.2%) | 17 | (20.0%) | 40 | (34.2%) |  |

Before PCV15: October 1, 2022-September 30, 2023; 1 year after PCV15: October 1, 2023-September 30, 2024; 2 years after PCV15: October 1, 2024-September 30, 2025.

Serotype 3 was the most prevalent serotype in all three periods but its proportion as a cause of IPD declined significantly when comparing the pre-PCV15 period (n= 39; 32.2%) with the first post-PCV15 (n=15; 17.6%; *p*= 0.01) and the second post-PCV15 (n= 22; 18.8%; *p*= 0.01) periods. The second most prevalent serotype along the three study periods was serotype 24F and its contribution to IPD was similar in the three study periods: pre-PCV15 (n=17; 14.0%), first post-PCV15 (n= 12; 14.1%; *p*= 0.9) and second post-PCV15 (n= 14; 12.0%; *p*= 0.6). Serotype 14, which was a residual serotype at baseline and during the first post-PCV15 period (one episode in each), emerged as the third most prevalent serotype in the second post-PCV15 period (n= 15; 12.8%; *p* < 0.001). (Figure 1)

**Figure 1:**
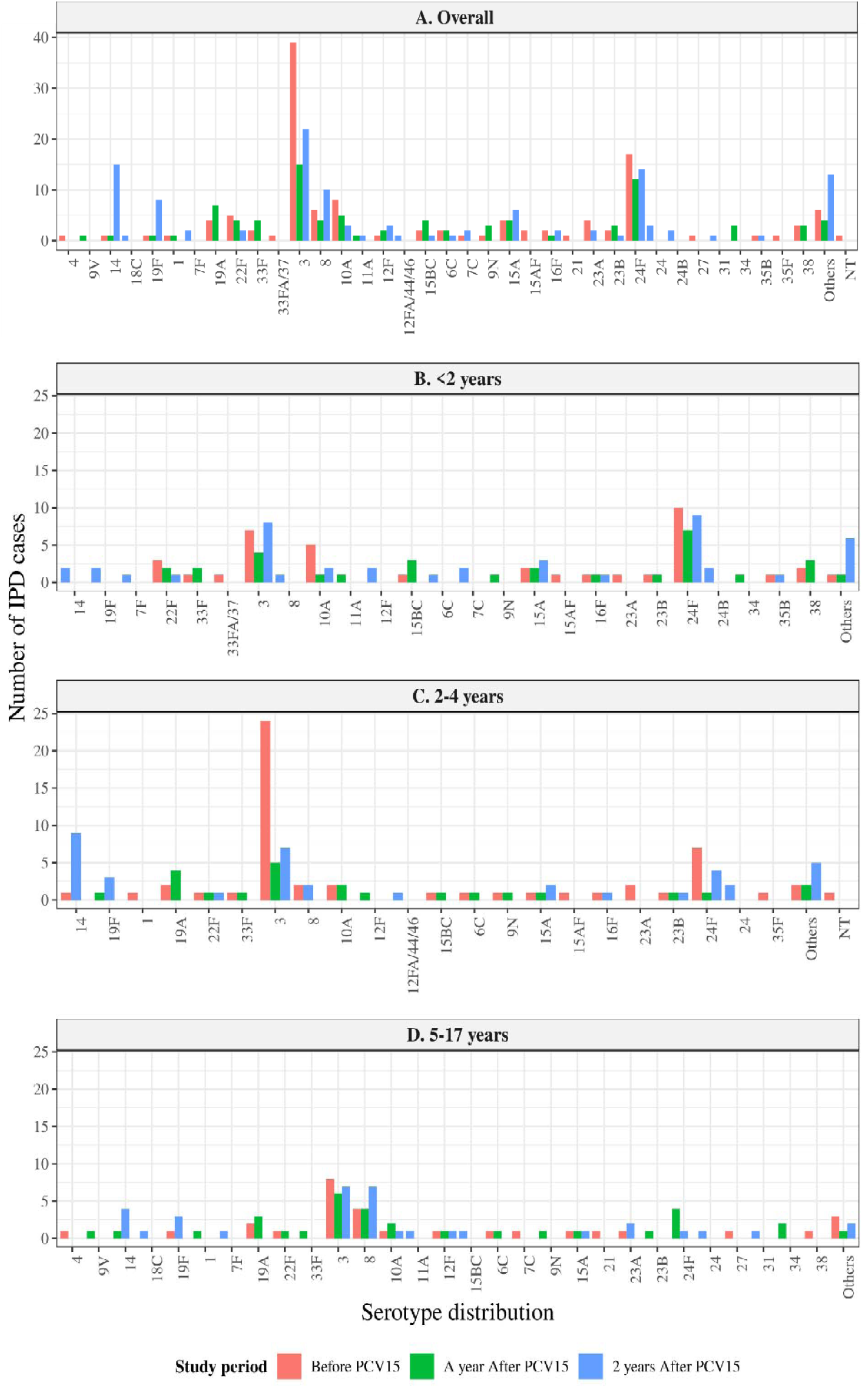
Pneumococcal serotypes distribution of invasive pneumococcal disease cases. Serotypes causing IPD across three age groups and overall before and after PCV15 implementation. The three periods analysed were: before PCV15 introduction (red bars), 1 year after PCV15 (green bars) and 2 years after PCV15 (blue bars).Others: serotypes that could not be classified due to low bacterial load. NT: Non-typable

Overall, a significant decrease in cumulative incidence of IPD was observed when comparing the pre-PCV15 vs the first post-PCV15 period (13.0 *vs.* 9.4 episodes/100,000 children< 18 years), representing a decrease of 28 % (OR: 0.72; 95% CI: 0.54-0.95; *p*= 0.02). However this protective effect was not sustained in the second year post-PCV15, when IPD cumulative incidence returned to baseline levels (OR: 1.00; 95%CI: 0.78-1.30; *p*=0.996).

The largest reduction in serotype-specific cumulative incidence was observed in IPD caused by serotype 3, which decreased from 4.1 to 1.6 episodes per 100,000 children under 18 years in the first post-PCV15 period, a decrease of 60% (OR: 0.40; 95% CI: 0.21-0.70; *p=*0.001) and to 2.3 episodes per 100,000 children under 18 years in the second period, a decrease of 42% (OR: 0.58; 95%CI: 0.34-0.98; *p*= 0.04). In contrast, a significant 690% increase in PCV7 covered serotypes was observed during the second post-PCV15period (OR: 7.90; 95% CI: 2.75-34.53; *p<*0.001), primarily driven by the increase of serotype 14. (Table 2 and Figure 1)

**Table 2:**
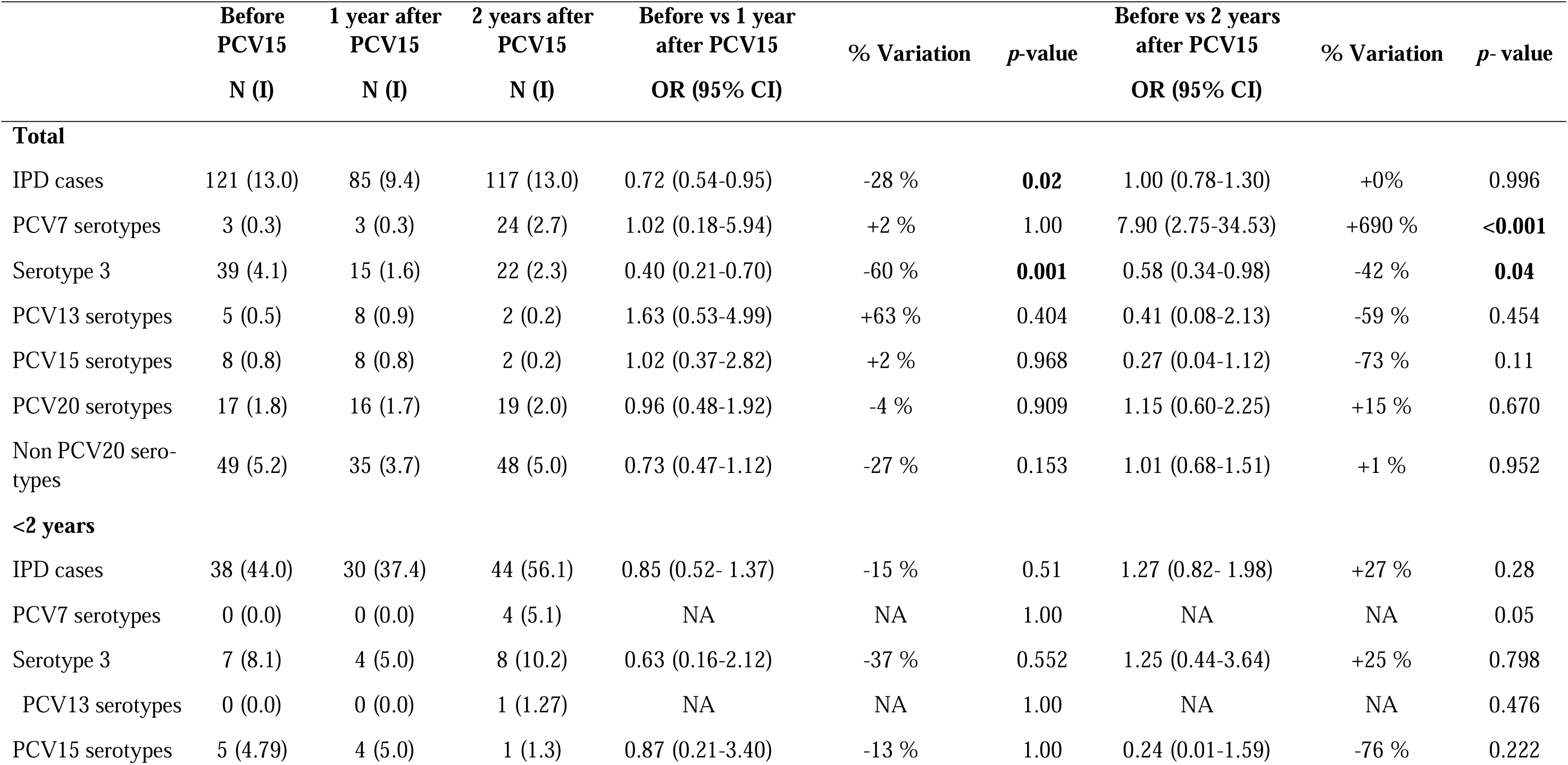
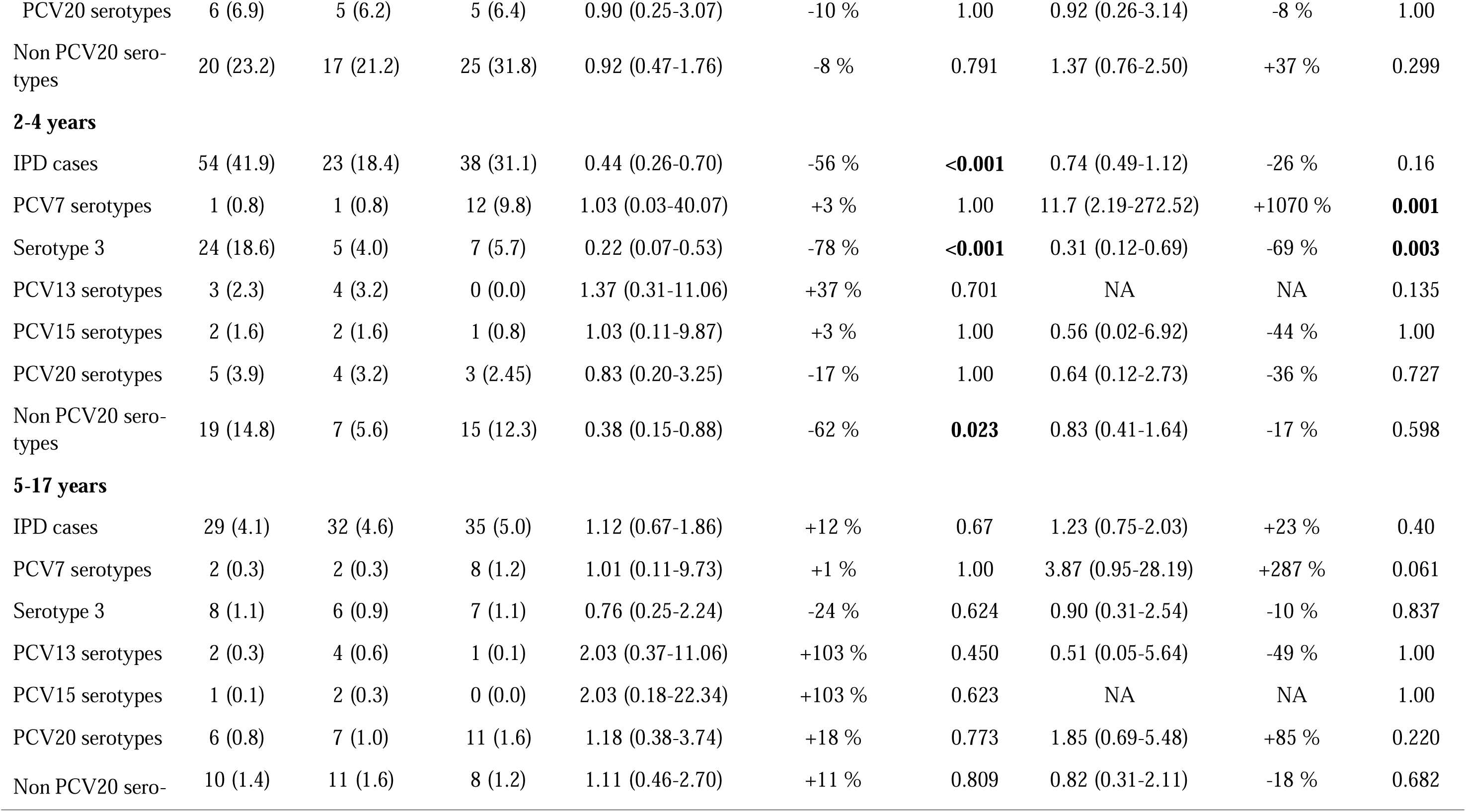
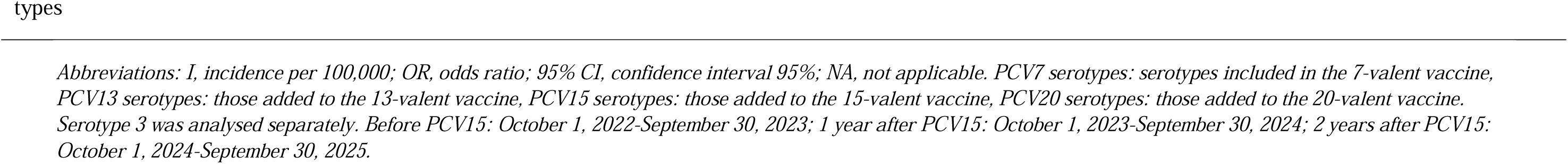
Incidence of Invasive Pneumococcal Disease before and after the introduction of PCV15 according to pneumococcal serotypes and age groups.

|  | Before<br>PCV15<br>N (I) | 1 year after<br>PCV15<br>N (I) | 2 years after<br>PCV15<br>N (I) | Before vs 1 year<br>after PCV15<br>OR (95% CI) | % Variation | *p*-value | Before vs 2 years<br>after PCV15<br>OR (95% CI) | % Variation | *p*- value |
| --- | --- | --- | --- | --- | --- | --- | --- | --- | --- |
| **Total** |  |  |  |  |  |  |  |  |  |
| IPD cases | 121 (13.0) | 85 (9.4) | 117 (13.0) | 0.72 (0.54-0.95) | -28 % | **0.02** | 1.00 (0.78-1.30) | +0% | 0.996 |
| PCV7 serotypes | 3 (0.3) | 3 (0.3) | 24 (2.7) | 1.02 (0.18-5.94) | +2 % | 1.00 | 7.90 (2.75-34.53) | +690 % | **<0.001** |
| Serotype 3 | 39 (4.1) | 15 (1.6) | 22 (2.3) | 0.40 (0.21-0.70) | -60 % | **0.001** | 0.58 (0.34-0.98) | -42 % | **0.04** |
| PCV13 serotypes | 5 (0.5) | 8 (0.9) | 2 (0.2) | 1.63 (0.53-4.99) | +63 % | 0.404 | 0.41 (0.08-2.13) | -59 % | 0.454 |
| PCV15 serotypes | 8 (0.8) | 8 (0.8) | 2 (0.2) | 1.02 (0.37-2.82) | +2 % | 0.968 | 0.27 (0.04-1.12) | -73 % | 0.11 |
| PCV20 serotypes | 17 (1.8) | 16 (1.7) | 19 (2.0) | 0.96 (0.48-1.92) | -4 % | 0.909 | 1.15 (0.60-2.25) | +15 % | 0.670 |
| Non PCV20 sero-<br>types | 49 (5.2) | 35 (3.7) | 48 (5.0) | 0.73 (0.47-1.12) | -27 % | 0.153 | 1.01 (0.68-1.51) | +1 % | 0.952 |
| **<2 years** |  |  |  |  |  |  |  |  |  |
| IPD cases | 38 (44.0) | 30 (37.4) | 44 (56.1) | 0.85 (0.52- 1.37) | -15 % | 0.51 | 1.27 (0.82- 1.98) | +27 % | 0.28 |
| PCV7 serotypes | 0 (0.0) | 0 (0.0) | 4 (5.1) | NA | NA | 1.00 | NA | NA | 0.05 |
| Serotype 3 | 7 (8.1) | 4 (5.0) | 8 (10.2) | 0.63 (0.16-2.12) | -37 % | 0.552 | 1.25 (0.44-3.64) | +25 % | 0.798 |
| PCV13 serotypes | 0 (0.0) | 0 (0.0) | 1 (1.27) | NA | NA | 1.00 | NA | NA | 0.476 |
| PCV15 serotypes | 5 (4.79) | 4 (5.0) | 1 (1.3) | 0.87 (0.21-3.40) | -13 % | 1.00 | 0.24 (0.01-1.59) | -76 % | 0.222 |
| PCV20 serotypes | 6 (6.9) | 5 (6.2) | 5 (6.4) | 0.90 (0.25-3.07) | -10 % | 1.00 | 0.92 (0.26-3.14) | -8 % | 1.00 |
| Non PCV20 sero-<br>types | 20 (23.2) | 17 (21.2) | 25 (31.8) | 0.92 (0.47-1.76) | -8 % | 0.791 | 1.37 (0.76-2.50) | +37 % | 0.299 |
| **2-4 years** |  |  |  |  |  |  |  |  |  |
| IPD cases | 54 (41.9) | 23 (18.4) | 38 (31.1) | 0.44 (0.26-0.70) | -56 % | **<0.001** | 0.74 (0.49-1.12) | -26 % | 0.16 |
| PCV7 serotypes | 1 (0.8) | 1 (0.8) | 12 (9.8) | 1.03 (0.03-40.07) | +3 % | 1.00 | 11.7 (2.19-272.52) | +1070 % | **0.001** |
| Serotype 3 | 24 (18.6) | 5 (4.0) | 7 (5.7) | 0.22 (0.07-0.53) | -78 % | **<0.001** | 0.31 (0.12-0.69) | -69 % | **0.003** |
| PCV13 serotypes | 3 (2.3) | 4 (3.2) | 0 (0.0) | 1.37 (0.31-11.06) | +37 % | 0.701 | NA | NA | 0.135 |
| PCV15 serotypes | 2 (1.6) | 2 (1.6) | 1 (0.8) | 1.03 (0.11-9.87) | +3 % | 1.00 | 0.56 (0.02-6.92) | -44 % | 1.00 |
| PCV20 serotypes | 5 (3.9) | 4 (3.2) | 3 (2.45) | 0.83 (0.20-3.25) | -17 % | 1.00 | 0.64 (0.12-2.73) | -36 % | 0.727 |
| Non PCV20 sero-<br>types | 19 (14.8) | 7 (5.6) | 15 (12.3) | 0.38 (0.15-0.88) | -62 % | **0.023** | 0.83 (0.41-1.64) | -17 % | 0.598 |
| **5-17 years** |  |  |  |  |  |  |  |  |  |
| IPD cases | 29 (4.1) | 32 (4.6) | 35 (5.0) | 1.12 (0.67-1.86) | +12 % | 0.67 | 1.23 (0.75-2.03) | +23 % | 0.40 |
| PCV7 serotypes | 2 (0.3) | 2 (0.3) | 8 (1.2) | 1.01 (0.11-9.73) | +1 % | 1.00 | 3.87 (0.95-28.19) | +287 % | 0.061 |
| Serotype 3 | 8 (1.1) | 6 (0.9) | 7 (1.1) | 0.76 (0.25-2.24) | -24 % | 0.624 | 0.90 (0.31-2.54) | -10 % | 0.837 |
| PCV13 serotypes | 2 (0.3) | 4 (0.6) | 1 (0.1) | 2.03 (0.37-11.06) | +103 % | 0.450 | 0.51 (0.05-5.64) | -49 % | 1.00 |
| PCV15 serotypes | 1 (0.1) | 2 (0.3) | 0 (0.0) | 2.03 (0.18-22.34) | +103 % | 0.623 | NA | NA | 1.00 |
| PCV20 serotypes | 6 (0.8) | 7 (1.0) | 11 (1.6) | 1.18 (0.38-3.74) | +18 % | 0.773 | 1.85 (0.69-5.48) | +85 % | 0.220 |
| Non PCV20 sero- | 10 (1.4) | 11 (1.6) | 8 (1.2) | 1.11 (0.46-2.70) | +11 % | 0.809 | 0.82 (0.31-2.11) | -18 % | 0.682 |

Abbreviations: *I*, incidence per 100,000; *OR*, odds ratio; 95% *CI*, confidence interval 95%; *NA*, not applicable. *PCV7* serotypes: serotypes included in the 7-valent vaccine, *PCV13* serotypes: those added to the 13-valent vaccine, *PCV15* serotypes: those added to the 15-valent vaccine, *PCV20* serotypes: those added to the 20-valent vaccine. Serotype 3 was analysed separately. Before *PCV15*: October 1, 2022-September 30, 2023; 1 year after *PCV15*: October 1, 2023-September 30, 2024; 2 years after *PCV15*: October 1, 2024-September 30, 2025.

The largest decline in IPD cumulative incidence was observed among children aged 2-4 years, with rates decreasing from 41.9 to 18.4 episodes per 100,000 children (OR: 0.44; 95% CI: 0.26–0.70; *p*<0.001) during the first post□PCV15 period; however, this reduction was not sustained overall, with cumulative incidence increasing to 31.1 episodes per 100,000 in the second period. This initial decline in this age group was largely driven by a marked reduction in serotype 3, which decreased from 18.6 to 4.0 episodes per 100,000 children in the first year after PCV15 introduction (78% reduction; OR: 0.22; 95% CI: 0.07-0.53; *p*<0.001) and remained significantly lower than baseline after two years (5.7 episodes per 100,000; 69% reduction; OR: 0.31; 95% CI: 0.12-0.69; *p*= 0.003). In contrast, in children aged 2-4 years, although a decrease in non□PCV20 serotypes was observed during the first post□PCV15 period from 14.8 to 5.6 episodes per 100,000 (OR: 0.38; 95% CI: 0.15-0.88; *p*= 0.023), this reduction was not sustained, with cumulative incidence returning to baseline levels in the second year (12.3 episodes per 100,000) (Table 2; Figure 1).

Sex-specific analyses showed divergent trends in IPD cumulative incidence following PCV15 introduction. Among male children, cumulative incidence declined significantly from 9.0 to 5.0 episodes per 100,000 in the first post□PCV15 year (OR: 0.55; 95% CI: 0.38-0.80; *p*= 0.001) and remained significantly lower after two years (5.8 episodes per 100,000; OR: 0.65; 95% CI: 0.46-0.91; *p*=0.013), with the greatest decrease observed in children aged 2-4 years. In contrast, female children experienced an increase in IPD cumulative incidence during the second post□PCV15 period, rising from 4.4 to 7.2 episodes per 100,000 (OR: 1.76; 95% CI: 1.19-2.66; *p*= 0.004), driven mainly by increases in those aged <2 years and 5–17 years (see table 3). By clinical presentation, a significant reduction in IPD was observed among cases presenting with pneumonia, particularly complicated pneumonia, during the first post□PCV15 period (OR: 0.47; 95% CI: 0.23-0.92; *p=*0.028). However, this decrease was not sustained in the second year following vaccine introduction (Table 3).

**Table 3:** Incidence of Invasive Pneumococcal Disease before and after the introduction of PCV15 according to sex, age groups and clinical features.

|  | Before<br>PCV15<br>N (I) | 1 year after<br>PCV15<br>N (I) | 2 years after<br>PCV15<br>N (I) | Before vs 1 year<br>after PCV15<br>OR (95% CI) | % Variation | p- value | Before vs 2 years<br>after PCV15<br>OR (95% CI) | % Variation | p- value |
| --- | --- | --- | --- | --- | --- | --- | --- | --- | --- |
| **Total** |  |  |  |  |  |  |  |  |  |
| Male | 83 (9.0) | 45 (5.0) | 52 (5.8) | 0.55 (0.38-0.80) | -45 % | **0.001** | 0.65 (0.46-0.91) | -35 % | **0.013** |
| Female | 38 (4.1) | 40 (4.4) | 65 (7.2) | 1.07 (0.69-1.69) | +7 % | 0.754 | 1.76 (1.19-2.66) | +76 % | **0.004** |
| Bacteriemia | 36 (3.8) | 25 (2.7) | 40 (4.3) | 0.71 (0.42-1.18) | -29 % | 0.187 | 1.15 (0.73-1.81) | +15 % | 0.549 |
| Pneumonia | 74 (7.8) | 46 (5.0) | 67 (7.2) | 0.64 (0.44-0.91) | -36% | **0.014** | 0.94 (0.67-1.30) | -6 % | 0.695 |
| *Non-complicated* | 48 (5.0) | 34 (3.7) | 50 (5.4) | 0.72 (0.46-1.12) | -28 % | 0.148 | 1.08 (0.72-1.60) | +8 % | 0.716 |
| *Complicated* | 26 (2.7) | 12 (1.3) | 17 (1.8) | 0.47 (0.23-0.92) | -53 % | **0.028** | 0.68 (0.36-1.24) | -32 % | 0.211 |
| Meningitis | 9 (1.0) | 12 (1.3) | 7 (0.8) | 1.35 (0.57-3.36) | +35 % | 0.494 | 0.81 (0.28-2.20) | -19 % | 0.676 |
| Others | 2 (0.2) | 2 (0.2) | 3 (0.3) | 1.02 (0.11-9.80) | +2 % | 1.000 | 1.51 (0.23-13.04) | +51 % | 0.683 |
| **<2 years** |  |  |  |  |  |  |  |  |  |
| Male | 27 (31.3) | 17 (21.2) | 18 (22.9) | 0.68 (0.36-1.24) | -32 % | 0.210 | 0.74 (0.40-1.33) | -26 % | 0.311 |
| Female | 11 (12.8) | 13 (16.2) | 26 (33.1) | 1.27 (0.56-2.92) | +27% | 0.564 | 2.57 (1.30-5.47) | +157 % | **0.006** |
| Bacteriemia | 10 (11.6) | 11 (13.7) | 19 (24.2) | 1.18 (0.49-2.87) | +18 % | 0.705 | 2.07 (0.98-4.68) | +107 % | 0.056 |
| Pneumonia | 24 (27.8) | 14 (17.4) | 21 (26.8) | 0.63 (0.31-1.21) | -37 % | 0.166 | 0.96 (0.53-1.73) | -4 % | 0.901 |
| *Non complicated* | 19 (22.0) | 12 (14.9) | 18 (22.9) | 0.68 (0.32-1.40) | -32 % | 0.299 | 1.04 (0.54-2.00) | +4 % | 1.00 |
| *Complicated* | 5 (5.8) | 2 (2.5) | 3 (3.8) | 0.45 (0.06-2.18) | -55 % | 0.455 | 0.67 (0.13-2.86) | -33 % | 0.729 |
| Meningitis | 3 (3.5) | 5 (6.2) | 3 (3.8) | 1.75 (0.41-9.11) | +75 % | 0.494 | 1.10 (0.19-6.40) | +10 % | 1.00 |
| Others | 1 (1.2) | 0 (0.0) | 1 (1.3) | NA | NA | 0.518 | 0.70 (0.04-11.27) | -30% | 0.827 |
| **2-4 years** |  |  |  |  |  |  |  |  |  |
| Male | 36 (28.0) | 10 (8.0) | 20 (16.4) | 0.29 (0.13-0.56) | -71 % | **<0.001** | 0.59 (0.33-1.00) | -41 % | 0.05 |
| Female | 18 (14.0) | 13 (10.4) | 18 (14.7) | 0.74 (0.35-1.52) | -26 % | 0.419 | 1.05 (0.54-2.04) | +5 % | 0.879 |
| Bacteriemia | 18 (14.0) | 4 (3.2) | 11 (9.0) | 0.23 (0.06-0.64) | -77 % | **0.004** | 0.64 (0.29-1.35) | -36 % | 0.27 |
| Pneumonia | 33 (25.3) | 14 (11.2) | 22 (18.0) | 0.44 (0.23-0.81) | -56 % | **0.007** | 0.70 (0.40-1.20) | -30% | 0.199 |
| *Non complicated* | 16 (12.4) | 10 (8.0) | 14 (11.4) | 0.65 (0.28-1.41) | -35 % | 0.277 | 0.922 (0.44-1.90) | -7.8 % | 0.826 |
| *Complicated* | 17 (13.2) | 4 (3.2) | 8 (6.5) | 0.25 (0.07-0.68) | -75 % | **0.007** | 0.50 (0.20-1.13) | -50 % | 0.09 |
| Meningitis | 3 (2.3) | 3 (2.4) | 3 (2.5) | 1.03 (0.18-5.98) | +3 % | 1.00 | 1.05 (0.18-6.12) | +5 % | 1.00 |
| Others | 0 (0.0) | 2 (1.6) | 2 (1.6) | NA | NA | 0.243 | NA | NA | 0.237 |
| **5-17 years** |  |  |  |  |  |  |  |  |  |
| Male | 20 (2.8) | 18 (2.6) | 14 (2.0) | 0.91 (0.48-1.74) | -9 % | 0.779 | 0.72 (0.35-1.42) | -28 % | 0.342 |
| Female | 9 (1.3) | 14 (2.0) | 21 (3.0) | 1.57 (0.68-3.80) | +57 % | 0.293 | 2.36 (1.11-5.48) | +136 % | **0.025** |
| Bacteriemia | 8 (1.1) | 10 (1.4) | 10 (1.4) | 1.26 (0.49-3.35) | +26 % | 0.628 | 1.27 (0.49-3.38) | +27 % | 0.614 |
| Pneumonia | 17 (2.4) | 18 (2.6) | 24 (3.4) | 1.07 (0.55-2.11) | +7% | 0.839 | 1.43 (0.78-2.73) | +43 % | 0.250 |
| *Non complicated* | 13 (1.8) | 12 (1.7) | 18 (2.6) | 0.94 (0.42-2.08) | -6 % | 0.869 | 1.41 (0.469-2.96) | +41 % | 0.345 |
| *Complicated* | 4 (0.6) | 6 (0.9) | 6 (0.9) | 1.50 (0.42-6.11) | +50 % | 0.546 | 1.51 (0.42-6.16) | +51 % | 0.544 |
| Meningitis | 3 (0.4) | 4 (0.6) | 1 (0.1) | 1.33 (0.28-7.24) | +33 % | 0.725 | 0.37 (0.01-3.20) | -63 % | 0.625 |
| Others | 1 (0.2) | 0 (0.0) | 0 (0.0) | NA | NA | 1.00 | NA | NA | 1.00 |

*Abbreviations: I, incidence per 100,000; OR, odds ratio; 95% CI, confidence interval 95%; NA, not applicable. Before PCV15: October 1, 2022-September 30, 2023; 1 year* *after PCV15: October 1, 2023-September 30, 2024; 2 years after PCV15: October 1, 2024-September 30, 2025.*

## DISCUSSION

We observed a significant decline in pediatric IPD caused by serotype 3 that persisted two years after PCV15 introduction into the universal childhood immunization program. However, this sustained serotype□specific reduction was offset in the second year by an increase in disease caused by PCV7 serotypes, resulting in no overall change in all□cause IPD cumulative incidence.

During the first year following PCV15 introduction, the marked serotype 3 decline, combined with a reduction in non□PCV20 serotypes among children aged 2-4 years, produced a significant overall IPD decrease. This early effect likely reflects indirect vaccine impact on nasopharyngeal carriage and transmission of vaccine□included serotypes. Additionally, temporal overlap between PCV15 implementation and the introduction of passive immunization with the monoclonal antibody nirsevimab against RSV in infants younger than 12 months warrants consideration [25]. RSV infection has been associated with nasopharyngeal microbiota dysbiosis and increaseded subsequent bacterial invasive disease risk [27,28]. Nevertheless, any RSV prevention contribution appears transient, as overall IPD cumulative incidence returned to baseline in the second year, despite the continued and significant reduction in serotype□3-specific IPD.

The sustained decrease of IPD caused by serotype 3 is encouraging because it has been one of the major serotypes causing IPD in Europe despite being included in the previously commercialized PCV13 vaccine [9–13]. These findings align with published data suggesting that PCV15 offers improved effectiveness against serotype 3 compared to both PCV13 and PCV20 vaccines [29].

The well□documented ability of PCVs to reduce nasopharyngeal carriage of vaccine serotypes is the principal mechanism underlying their indirect, or herd, effects [30] . By preventing colonization in vaccinated individuals, PCVs limit onward transmission of *Streptococcus pneumoniae*, thereby conferring protection to unvaccinated populations. Early infancy represents a critical window for pneumococcal acquisition, as primary colonization typically occurs within the first year of life and has been reported in nearly all children by 12 months of age [31]. Consequently, vaccination during this period may prevent initial colonization events and substantially disrupt the primary reservoir for pneumococcal transmission. This mechanism provides a biologically plausible explanation for the marked reductions in IPD observed as early as the first year after PCV15 introduction, with declines exceeding vaccine coverage and extending to unvaccinated age groups. Similar early and disproportionate indirect effects were previously reported following the introduction of PCV7 [6] and PCV13 [9].

Contrary to the expectation that serotype replacement would be driven predominantly by non□vaccine serotypes [8,32], the second post-PCV15 year was characterized by a marked increase in serotype 14 disease. This, vaccine serotype was included in previous formulations (PCV7 and PCV13) and was present at residual levels during the baseline period. This finding suggests that post-vaccination serotype dynamics may be more complex than classical replacement by non-vaccine serotypes alone. One possible explanation is reduced immunogenicity associated with higher□valent pneumococcal conjugate vaccines, which has been proposed to attenuate immune responses to certain shared serotypes [33,34]. In hight-valent formulations, competition for immune processing or carrier-induced epitope suppression could theoretically reduce protection against previously well□controlled serotypes, allowing their re□emergence despite ongoing inclusion in the formulation [35]. Although the underlying mechanisms remain to be fully elucidated, our findings highlight the need for close surveillance of individual vaccine serotypes following the introduction of higher□valent PCVs.

An additional contextual factor that warrants consideration is the concurrent use of higher□valent pneumococcal conjugate vaccines in other populations during the period when serotype 14 was observed to increase. At that time, vaccines with an even higher valency were being administered to adults in Catalonia and to pediatric populations in other regions of the country, potentially altering pneumococcal transmission dynamics at a broader population level (21,24). Heterogeneous vaccine pressures across age groups and geographic areas may have contributed to the increased circulation of specific serotypes, including serotype 14, through mechanisms that are not fully understood by localized pediatric vaccination programs. This context highlights the complexity of pneumococcal ecology in settings with overlapping vaccination strategies and further underscores the need for integrated surveillance across age groups and regions.

Most large surveillance studies have consistently reported higher IPD incidence among male than female children, including during the PCV7 and PCV13 eras [36]. However, our post□PCV15 data show a relative increase among girls, particularly in those aged <2 years and 5–17 years. This increase may reflect age□specific serotype replacement, local contact patterns, and random variation, rather than reversal of the known male excess. Global analyses do not suggest intrinsic serotype invasiveness differences by sex [3,37]. However, our findings are consistent with observations from the Netherlands following PCV7 and PCV10 introduction, where overall IPD incidence declined in men aged 20-39 years but transiently increased in woman, driven by replacement disease [38]. Together, these observations indicate that sex□specific IPD patterns can emerge following PCV introduction and support continued surveillance stratified by sex and age to better elucidate the underlying biological and epidemiological mechanisms

Our study has several limitations. First, surveillance was limited to two years following PCV15 implementation. Although the long□term impact of PCV15 in Catalonia will require continued monitoring, these data provide an important early assessment of vaccine impact and generate hypotheses relevant to future vaccination policy decisions. Notably, similar early IPD decline patterns have been reported following the introduction of previous pneumococcal conjugate vaccines, including PCV7 and PCV13 [6,39], and these patterns were instrumental in supporting their subsequent incorporation into immunization programs.

Second, we did not formally assess nirsevimab’s potential impact on IPD cumulative incidence. Nirsevimab introduction overlapped temporally with PCV15 implementation, and RSV infection has been associated with nasopharyngeal environment alterations that may lead to bacterial infections [40]. However, this potential confounding effect is unlikely to fully account for our findings. Overall IPD reduction was restricted to the first post□PCV15 year, whereas the decline in vaccine serotype 3□specific IPD remained significant in the second year, when no sustained overall IPD reduction was observed. This temporal dissociation suggests that the effect on serotype 3 is vaccine□mediated, rather than indirectly attributable to RSV prevention.

Third, not all participating hospitals had routine access to pneumococcal real□time PCR for the microbiological diagnosis of IPD, which may have led to an underestimation of the true disease burden. However, this limitation is unlikely to have biased the observed trends, as diagnostic practices remained stable throughout the study period and this potential under□ascertainment would be expected to apply similarly across all periods, thus preserving the validity of the comparative analyses

In conclusion, PCV15 introduction was associated with a sustained and significant reduction in serotype 3 IPD over a two□year period. However, this serotype□specific benefit was offset by a marked increase in disease caused by PCV7 serotypes, resulting in no net reduction in overall IPD cumulative incidence. These findings highlight both the early impact and the complex serotype dynamics following PCV15 implementation, underscoring the need for continued serotype□specific surveillance.

## Data Availability

All data produced in the present work are contained in the manuscript

## FUNDING

This work was supported by the ISCIII Sub-Directorate General for Evaluation and Promotion of Research (Projects CMA: PI23/00049, MFDS: PI23/00051) cofounded by Fondo Europeo de Desarrollo Regional (FEDER) and Agència de Gestió d’Ajuts Universitaris i de Recerca (AGAUR) (Grant CMA: 2021 SGR 00396 and AD: 2021/SGR 0072). MC is supported by a PFIS fellowship FI 24/00206 and RV is supported by ARISTOS that has received funding from the European Union’s Horizon Europe research and innovation program under the Marie Sklodowska-Curie grant agreement No 101081334. The funding sources had not role in the writing up of the manuscript and in the decision to submit for publication.

## ACKNOWLEDGEMENTS

We would like to thank the rest of members of the Catalan Study Group of invasive pneumococcal disease: M. Fernández, C. Pitart, and M. Espasa, Hospital Clínic Barcelona;; E. Folch and P. Ortiz, Clínica Girona; E. Capdevila and P. Hernandez, Consorci del Maresme i la Selva Calella-Blanes; ; S. Gonzalez, Hospital Dos de Maig,CLILAB Diagnòstics; C. Mora and M. Iglesias, Hospital de Figueres; A. Pulido and M. Armas, Hospital de Granollers; R.K. Santos da Silva, Hospital Esperit Sant; R. Clivillé, Hospital de Sant Joan Despí Moisès Broggi, CLILAB Diagnòstics; A. Soler and S. Hernandez, Hospital Sant Joan de Deu de Barcelona; A. Garcia, Hospital d’Igualada,CLILAB Diagnòstics; J. Lucena, Hospital General de l’Hospitalet, CLILAB Diagnòstics;; E. Sanfeliu, Hospital Sant Jaume d’Olot; N. Torrellas, Hospital de Palamós; A. González-Cuevas, Hospital Sant Joan de Deu de Sant Boi; C. Esteban, Hospital Residència Sant Camil, CLILAB Diagnòstics ; X. Clivillé and S. Noguer, Hospital Sant Pau I Santa Tecla; S. Noguer, Hospital del Vendrell; MO Pérez, Hospital Verge de la Cinta;.

## CONTRIBUTIONS

Carmen Muñoz-Almagro, designed the study, funding acquisition, analyzed data, wrote the paper, supervised the study

Maria Cisneros, analyzed data, performed experiments, wrote the paper

Claudia Alcaraz, collected data

Sonia Broner, collected data

Fernando Moraga, collected data

Anna Rosell, collected data

Alvaro Díaz, collected data

Desiree Henares, collected data, performed experiments

Gerard Gonzalez-Comino, collected data, performed experiments

Belen Viñado, collected data, performed experiments

Frederic Gómez-Bartomeu, collected data, performed experiments

Clara Marco, collected data

Sebastian Gonzalez-Peris, collected data

Jaume Llaberia, collected data

Jessica Galvez, collected data

Amaresh Perez-Argüello, collected data, performed experiments

Rosauro Varo, collected data

Jordi Iglesies, collected data

Cristina Esteva, collected data

Mayuli Armas, collected data

Conchita Izquierdo, collected data

Miguel Blanco-Fuertes, collected data

Nuria Torrellas, collected data

Mar Olga Perez, collected data

Ines T Valle, collected data

Marian Navarro, collected data

Alba Rivera, collected data

Montserrat Colomer, collected data

Laura Solaz, collected data

Miquel Mico, collected data,

Juan J Garcia, collected data, funding acquisition

Angela Dominguez, collected data, funding acquisition

Pilar Ciruela, collected data, funding acquisition, supervised the study

Mariona F de Sevilla, collected data, funding acquisition, supervised the study

All authors and Catalan Study Group of Invasive Pneumococcal disease discussed the results and critically reviewed, discussed and accepted the final version of the manuscript.

## CONFLICT OF INTERESTS STATEMENT

Carmen Muñoz-Almagro reports travel grants from Pfizer, MSD, and personal fees as speaker from Sanofi-Pasteur, Pfizer and MSD. Alvaro Diaz report travel grant from Pfizer, Mariona F de Sevilla report personal fees as speaker from Pfizer and MSD. The rest of authors declare no conflicts of interest.

## Notes

### Competing Interest Statement

Carmen Munoz-Almagro reports travel grants from Pfizer, MSD, and personal fees as speaker from Sanofi-Pasteur, Pfizer and MSD. Alvaro Diaz report travel grant from Pfizer, Mariona F de Sevilla report personal fees as speaker from Pfizer and MSD. The rest of authors declare no conflicts of interest.

### Summary of Updates

This version of the manuscript has been revised to correct a typographical error in one of the study denominators, which resulted in a non significant alteration of some of the incidence calculations.

