## Supplementary material for "CHANGES IN INCIDENCE AND SEROTYPE DISTRIBUTION OF PEDIATRIC INVASIVE PNEUMOCOCCAL DISEASE AFTER THE INTRODUCTION OF 15-VALENT PNEUMOCOCCAL CONJUGATE VACCINE IN CATALONIA, SPAIN. A MULTICENTER SURVEILLANCE STUDY"

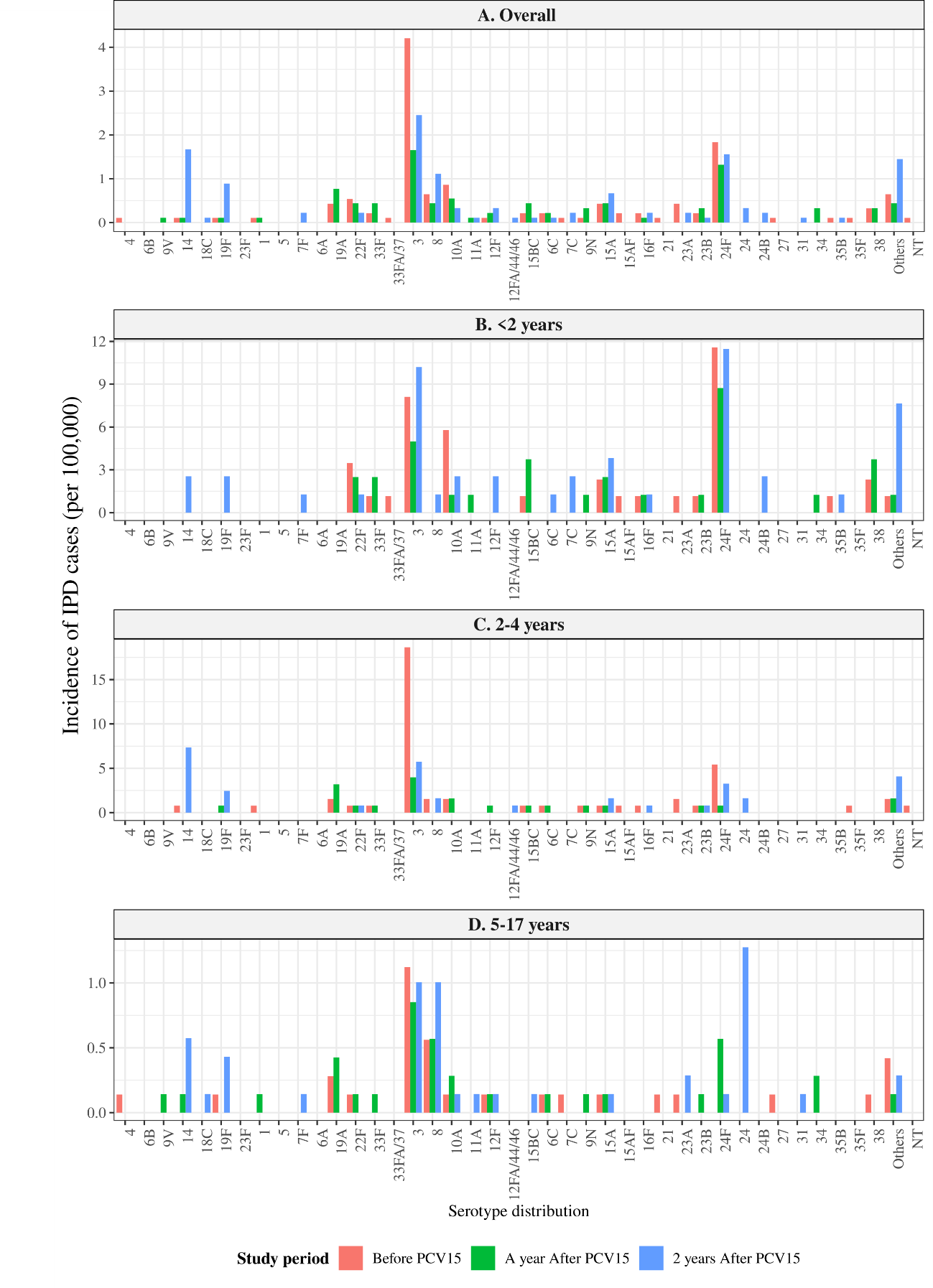
**Figure S1: Pneumococcal serotypes distribution and incidence per 100.000 person-year of invasive pneumococcal disease cases.**

Serotypes causing IPD across three age groups and overall before and after PCV15 implementation. The three periods analysed were: before PCV15 introduction (red bars), 1 year after PCV15 (green bars) and 2 years after PCV15 (blue bars). Others: serotypes that could not be classified due to low bacterial load. NT: Non-typable

**Table S1: List of Hospitals Contributing Samples to the Study**

|  | Hospital/Institution |
| --- | --- |
| 1 | Hospital Vilafranca - Alt Penedès- CLILAB Diagnòstics |
| 2 | Hospital de Barcelona |
| 3 | Hospital Clínic |
| 4 | Clínica Girona |
| 5 | Hospital de Sant Jaume de Calella - Consorci del Maresme i la Selva |
| 6 | Hospital Comarcal de Blanes- Consorci del Maresme i la Selva |
| 7 | Hospital Dos de Maig- CLILAB Diagnòstics |
| 8 | Hospital de Figueres |
| 9 | Hospital Sant Joan de Déu de Manresa - Fundació Althaia |
| 10 | Clínica Sant Josep- Fundació Althaia |
| 11 | Hospital de Granollers |
| 12 | Hospital Esperit Sant |
| 13 | Hospital de Nens |
| 14 | Hospital Sant Joan Despí Moisès Broggi- CLILAB Diagnòstics |
| 15 | Hospital Dr. Josep Trueta - Laboratori Clínic Territorial |
| 16 | Hospital de Campdevanol- Laboratori Clínic Territorial |
| 17 | Hospital Santa Caterina- Laboratori Clínic Territorial |
| 18 | Hospital General de Catalunya |
| 19 | Hospital de la Santa Creu i Sant Pau |
| 20 | Hospital Sant Joan de Déu |
| 21 | Hospital de Igualada- CLILAB Diagnòstics |
| 22 | Hospital Joan XXIII Tarragona |
| 23 | Hospital General de l'Hospitalet - CLILAB Diagnòstics |
| 24 | Hospital de Mataró |
| 25 | Hospital Olot |
| 26 | Hospital de Palamós |
| 27 | Hospital de Reus |
| 28 | Hospital de Sant Boi |
| 29 | Hospital Residencia Sant Camil |
| 30 | Hospital Santt Pau i Santa Tecla |
| 31 | Hospital del Vendrell |
| 32 | Hospital Verge de la Cinta Tortosa |
| 33 | Hospital Vall Hebrón |
| 34 | Hospital de Vic |

**Table S2: Children population in surveillance in 34 Hospitals of Catalonia by year**

|  | Before PCV15 | 1 year after PCV15 | 2 years after PCV15 |
| --- | --- | --- | --- |
| Children 0-<2 years | 86,306 | 80,202 | 78,466 |
| Children 2-< 5 years | 128,773 | 125,339 | 122,345 |
| Children 5-< 18 years | 712,686 | 703,695 | 696,934 |
| Children <18 years | 927,766 | 909,236 | 897,747 |

Before PCV15: October 1, 2022-September 30, 2023; 1 year after PCV15: October 1, 2023-September 30, 2024; 2 years after PCV15: October 1, 2024-September 30, 2025.
